# Closing the health inequity gap during the pandemic: COVID-19 mortality among racial and ethnic groups in Connecticut, March 2020 to December 2021

**DOI:** 10.1101/2022.02.18.22270820

**Authors:** Olivia Schultes, Margaret L. Lind, Jessica Brockmeyer, Peri Sosensky, Derek A.T. Cummings, Albert I. Ko

## Abstract

COVID-19 has disproportionally burdened racial and ethnic minority groups within the United States. Leveraging statewide data, we examined the evolution of racial and ethnic disparities in COVID-19 related deaths among Connecticut residents residing in non-congregate settings over three periods of the COVID-19 pandemic. Despite observing large disparities in the age-adjusted mortality rates between Hispanics, non-Hispanic Blacks, and non-Hispanic Whites during the initial pandemic period (March to August 2020), we observed meaningful reductions in the disparities during the subsequent periods (August 2020 to July 2021; July to mid December 2021). Further, during the third period, we failed to find a significant difference in age-adjusted mortality between non-Hispanic Blacks and non-Hispanic Whites. These findings provide evidence that attenuation of racial and ethnic disparities in COVID-19 related outcomes are achievable.

## Introduction

Significant racial and ethnic disparities in COVID-19 outcomes have been documented in the US.^1–5^ However, state and national level findings have been mostly based on aggregated data of populations residing in congregate and non-congregate settings.^1–5^ This approach may underestimate outcome disparities in community settings, especially in states that experienced a large COVID-19 burden in nursing homes.^5^ Herein, we identified COVID-19-related deaths associated with congregate and non-congregate residential settings in Connecticut and characterized the evolution of racial and ethnic disparities in COVID-19-related mortality among populations residing in non-congregate settings.

## Methods

We retrospectively identified COVID-19-related deaths reported between March 1^st^ 2020 and December 13^th^ 2021 from the Connecticut Electronic Disease Surveillance System and determined residential status according to recorded address (*Supplement*).^5,6^ For the population residing in non-congregate settings, we estimated age-adjusted mortality for composite race and ethnicity categories (Hispanic, non-Hispanic Black and non-Hispanic White) using 2019 census estimates (*Supplement*).^5^ We estimated age-adjusted mortality rate ratios (MRR) for Hispanic and non-Hispanic Blacks, relative to non-Hispanic Whites, for three periods (March 1, 2020 – August 25, 2020, August 26, 2020 – July 12, 2021 and July 13, 2021 – December 13, 2021; Figure.C; *Supplement*), each of which encompassed an epidemic wave, and tested for changes in MRR using a Poisson Regression. This research was deemed *Not Human Subjects Research* by the Yale Institutional Review Board.

**Figure.**
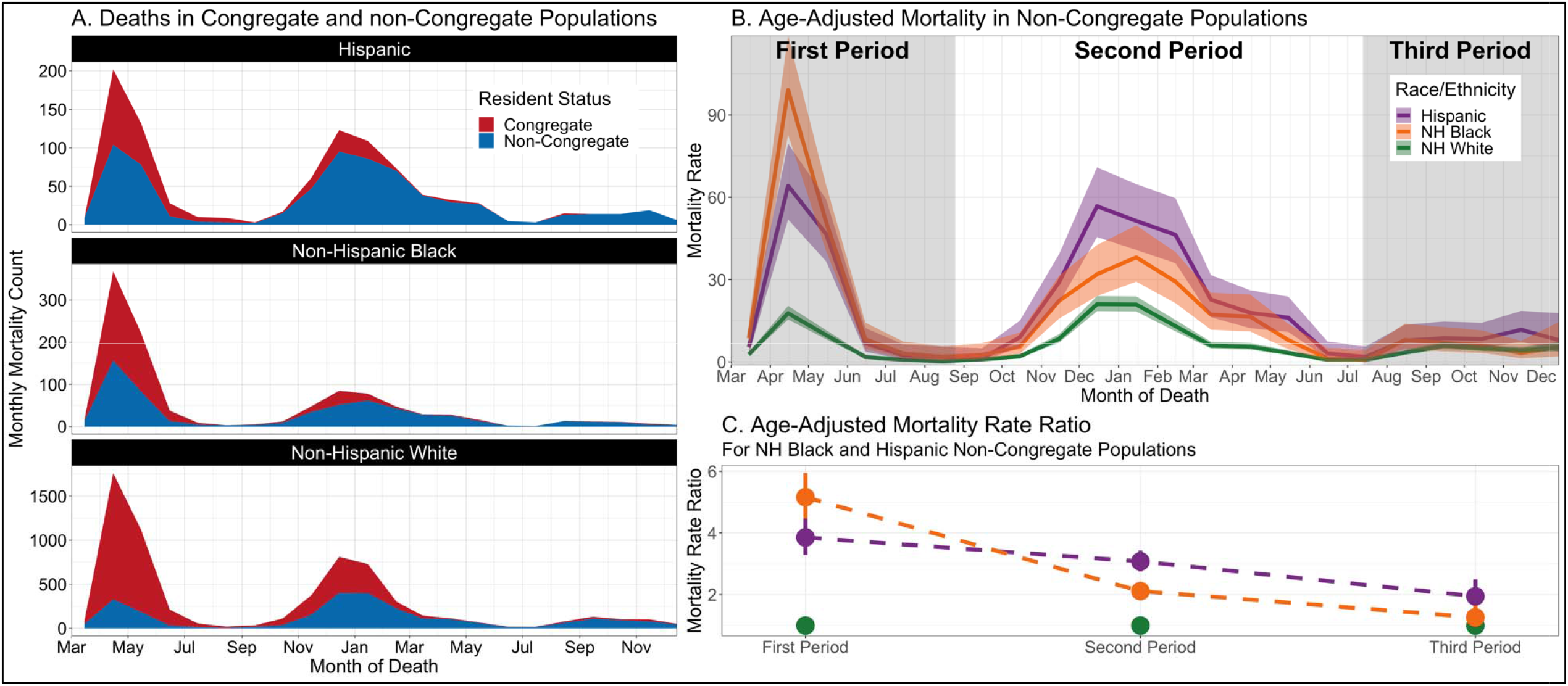
Temporal Trends in COVID-19-Related Deaths and Mortality among the Connecticut Population Residing in Congregate and Non-Congregate Settings, according to Composite Race and Ethnic Groups, March 2020 to December 2021. Panel A: Monthly COVID-19 deaths by race/ethnicity and residency status; Panel B: Age-adjusted monthly COVID-19 mortality (deaths per 100,000 population-months) by composite race and ethnic groups in the population residing in non-congregate settings; Panel C: Ratios comparing mortality among non-Hispanic Black (NH Black) and Hispanic to non-Hispanic White (NH White) populations residing in non-congregate settings during each of the three epidemic periods. Congregate and non-congregate residency status was defined based on recorded residential address at the SARS-CoV-2 infection was reported.

## Results

During the study period, 8,446 COVID-19-related deaths were reported among Hispanic, non-Hispanic Black and non-Hispanic White populations in Connecticut, of which 4,630 (54.8%) and 3,816 (45.2%) occurred among residents of congregate and non-congregate settings, respectively. Congregate facility-associated deaths accounted for 74.6% (3205/4299), 37.7% (1341/3557) and 14.2% (84/590) of deaths in the 1^st^, 2^nd^, and 3^rd^ periods, respectively (Figure.A). Among congregate facility-associated deaths, 5.6% (259), 10.0% (463) and 84.4% (3,908) were Hispanic, non-Hispanic Black and non-Hispanic White, respectively. The characteristics of deaths among residents of non-congregate settings are shown in the Table.

**Table.** Age-Adjusted COVID-19 Mortality and Mortality Rate Ratios in the Connecticut Non-Congregate Population, According to Composite Race and Ethnicity Groups, During Three Pandemic Periods.

| Group/Period<sup>b</sup> | Population-Months of Observation<sup>a</sup> | No. Deaths | Crude Mortality Rate<sup>c</sup> | Age-Adjusted Mortality (95% CI)<sup>c,d</sup> | MRR (95% CI)<sup>d</sup> | MRR normalized to 1<sup>st</sup> Wave (95% CI)<sup>d</sup> |
| --- | --- | --- | --- | --- | --- | --- |
| **Non-Hispanic White** | 54,808,226 | 2532 | 4.6 | 6.4 (5.8, 7.0) | (ref) | - |
| 1<sup>st</sup> period | 14,940,068 | 613 | 4.1 | 5.7 (5.1, 6.4) | (ref) | - |
| 2<sup>nd</sup> period | 26,942,482 | 1524 | 5.7 | 7.8 (7.1, 8.6) | (ref) | - |
| 3<sup>rd</sup> period | 12,925,676 | 395 | 3.1 | 4.2 (3.7, 4.8) | (ref) | - |
| **Non-Hispanic Black** | 8,601,736 | 592 | 6.9 | 17.3 (15.4, 19.5) | 2.7 (2.5, 3.0) | - |
| 1<sup>st</sup> period | 2,344,730 | 272 | 11.6 | 29.3 (25.3, 34.0) | 5.2 (4.5, 5.9) | (ref) |
| 2<sup>nd</sup> period | 4,228,418 | 277 | 6.6 | 16.5 (14.2, 19.1) | 2.1 (1.9, 2.4) | 0.4 (0.3, 0.5) |
| 3<sup>rd</sup> period | 2,028,587 | 43 | 2.1 | 5.3 (3.9, 7.3) | 1.3 (0.9, 1.7) | 0.2 (0.2, 0.3) |
| **Hispanic** | 14,015,129 | 692 | 4.9 | 19.6 (17.5, 22.1) | 3.1 (2.8, 3.4) | - |
| 1<sup>st</sup> period | 3,820,357 | 209 | 5.5 | 21.9 (18.6, 25.8) | 3.9 (3.3, 4.5) | (ref) |
| 2<sup>nd</sup> period | 6,889,520 | 415 | 6.0 | 24.0 (21.1, 27.4) | 3.1 (2.8, 3.4) | 0.8 (0.7, 1.0) |
| 3<sup>rd</sup> period | 3,305,252 | 68 | 2.1 | 8.2 (6.3, 10.6) | 1.9 (1.5, 2.5) | 0.5 (0.4, 0.7) |

<sup>a</sup>Person months estimated as the state population times the number of months included in each wave, partial months were estimated using an average month duration of 30.5 days
<sup>b</sup>Periods defined using the low points between three large waves in COVID-19 deaths in Connecticut (1<sup>st</sup> period: March 1, 2020 to August 25, 2020; 2<sup>nd</sup> period: August 26, 2020 to July 12, 2021; 3<sup>rd</sup> period: July 13, 2021 to December 13, 2021 (date of data collection))
<sup>c</sup>Crude and age-adjusted rates presented in events per 100,000 population months
<sup>d</sup>MRR: Mortality Rate Ratio; CI: 95% Confidence Interval

Overall, age-adjusted mortality among residents of non-congregate settings was highest for Hispanics (19.6/100,000 population-months), followed by non-Hispanic Blacks (17.3/100,000 population-months) and non-Hispanic Whites (6.4/100,000 population-months). During the 1^st^ period, non-Hispanic Blacks had a 5.2 (95% confidence interval [CI], 4.5-5.9) times higher mortality than non-Hispanic Whites. However, during the 3^rd^ period, the disparity decreased by 80% and mortality between non-Hispanic Blacks and non-Hispanic Whites did not differ significantly (MRR: 1.3 [0.9-1.7]).

Although Hispanics had a lower MRR (3.9 [3.3-4.5]) than non-Hispanic Blacks during the 1^st^ period, they had higher MRR during the 2^nd^ and 3^rd^ periods. Mortality for Hispanics remained significantly higher than for non-Hispanic Whites during the 3^rd^ period (MRR 1.9 [1.5-2.5]; Figure/Table).

## Discussion

During the initial pandemic period, we found COVID-19 mortality to be nearly five and four times greater for non-Hispanic Black and Hispanic than non-Hispanic White Connecticut residents residing in non-congregate settings. The high magnitude of these disparities compared with those reported during similar periods from other US regions (MRR 1.2-4.0)^1,2,4,5^ indicates that the disaggregation of congregate-facility events may reveal levels of disparity not previously appreciated.

Our findings suggest that attenuation of racial and ethnic disparities is an achievable public health goal. We observed a decline in disparities in COVID-19-related mortality, as found in prior studies.^1,3^. While we found no significant difference in the COVID-19-related mortality between non-Hispanic Blacks and non-Hispanic Whites during the 3^rd^ pandemic period, significant disparities remained between Hispanics and non-Hispanic Whites.

The limitations of our analysis include the use of a COVID-19-related death endpoint and the potential for misclassification of race, ethnicity and residency status. Further investigations are needed to disentangle the role interventions, such as social protection programs, testing and vaccination, had on reducing disparities in COVID-19 mortality, which may inform effective, generalizable policies targeting the health equity challenge posed by the pandemic.

## Data Availability

All data produced in the presented study are available upon reasonable request to the authors.

## Acknowledgments

The authors thank Charles Powell for accessing the data. Matthew Cartter, Lynn Sosa, and Karyn Backus for helpful comments on the manuscript.

## Supplemental Methods

### Methods

#### Population Denominators

We used total Connecticut population data from the 2019 US Census estimates then calculated age-adjusted rates for composite race and ethnic groups among non-congregate residence settings.^1^ We did not subtract counts for the population residing in congregate facilities from the denominators, since age, race and ethnicity stratified data is not available for this population. However, these estimates of age-adjusted rates are a reasonable approximation since congregate residential populations account for approximately 1% of the Connecticut population.

1. US Census Bureau. Annual County Resident Population Estimates by Age, Sex, Race, and Hispanic Origin: April 1, 2010 to July 1, 2019 (CC-EST2019-ALLDATA). Accessed February 7, 2022. https://www.census.gov/data/tables/time-series/demo/popest/2010s-counties-detail.html

#### Pandemic Period Definition

We defined pandemic periods based on the lowest 7-day rolling average of daily deaths (nadir) between three large COVID-19 epidemic waves in Connecticut. The three waves occurred in the spring of 2020, fall of 2020 through spring of 2021, and fall of 2021 through the end of the study period (December 13, 2021). The two nadirs were on August 25^th^, 2020, and July 12^th^, 2021, corresponding with 7-day rolling average death counts of 1.13 and 1.29, respectively.

#### Ascertainment of COVID-19 related deaths in congregate and non-congregate settings

Connecticut Department of Public Health (DPH) defines congregate residential settings as long-term care facilities (LTCFs), assisted living facilities, and correctional facilities. At the beginning of the pandemic, COVID-19 cases and deaths were defined by DPH as congregate setting residents if the geocoded residential address extracted from the Connecticut Electronic Disease Surveillance System fell within 150 feet of a registered congregate setting. As of February 2021, DPH used address-based text-matching to registered congregate settings as the method of identifying COVID-19 cases and deaths in congregate facility residences.

